# Acylcarnitines metabolism in depression: association with diagnostic status, depression severity and symptom profile in the NESDA cohort

**DOI:** 10.1101/2024.02.14.24302813

**Authors:** Silvia Montanari, Rick Jansen, Daniela Schranner, Gabi Kastenmüller, Matthias Arnold, Delfina Janiri, Gabriele Sani, Sudeepa Bhattacharyya, Siamak Mahmoudian Dehkordi, Boadie W Dunlop, A. John Rush, Brenda W. H. J. Penninx, Rima Kaddurah-Daouk, Yuri Milaneschi

## Abstract

**Background:** Acylcarnitines (ACs) are involved in bioenergetics processes that may play a role in the pathophysiology of depression. Studies linking AC levels to depression are few and provide mixed findings. We examined the association of circulating ACs levels with Major Depressive Disorder (MDD) diagnosis, overall depression severity and specific symptom profiles.

**Methods:** The sample from the Netherlands Study of Depression and Anxiety included participants with current (n=1035) or remitted (n=739) MDD and healthy controls (n=800). Plasma levels of four ACs (short-chain: acetylcarnitine C2 and propionylcarnitine C3; medium-chain: octanoylcarnitine C8 and decanoylcarnitine C10) were measured. Overall depression severity as well as atypical/energy-related (AES), anhedonic and melancholic symptom profiles were derived from the Inventory of Depressive Symptomatology.

**Results:** As compared to healthy controls, subjects with current or remitted MDD presented similarly lower mean C2 levels (Cohen’s d=0.2, p≤1e-4). Higher overall depression severity was significantly associated with higher C3 levels (ß=0.06, SE=0.02, p=1.21e-3). No associations were found for C8 and C10. Focusing on symptom profiles, only higher AES scores were linked to lower C2 (ß=-0.05, SE=0.02, p=1.85e-2) and higher C3 (ß=0.08, SE=0.02, p=3.41e-5) levels. Results were confirmed in analyses pooling data with an additional internal replication sample from the same subjects measured at 6-year follow-up (totaling 4195 observations).

**Conclusions:** Small alterations in levels of short-chain acylcarnitine levels were related to the presence and severity of depression, especially for symptoms reflecting altered energy homeostasis. Cellular metabolic dysfunctions may represent a key pathway in depression pathophysiology potentially accessible through AC metabolism.

## INTRODUCTION

Depression is the second-leading cause of disability worldwide^[1]^. The detrimental impact of depression includes sequelae that extend beyond mental health, including increased the risk for the development of cardiometabolic conditions such as cardiovascular disease and diabetes. Immuno-metabolic dysregulations have been proposed as mechanisms contributing to the overlapping pathophysiology of depression and cardiometabolic disorders ^[2]^.

Mitochondrial dysfunction has been recently proposed as a key pathophysiological mechanism^[3]^ linked to processes commonly found in depression, including neurotoxicity, impaired neuroplasticity, inflammation and insulin resistance^[2–5]^. Emerging evidence suggests a potential association of altered levels of acylcarnitines (ACs), which are involved in mitochondrial fatty acids β-oxidation, with insulin resistance, cardiovascular and neurodegenerative diseases ^[6–9]^. Mixed findings on altered ACs levels in depression have recently emerged from small clinical samples of subjects with Major Depressive Disorder (MDD)^[10,11]^ or larger samples from the general population^[12]^ with measures of depression limited to self-report symptom questionnaires. In a previous study^[13]^, we leveraged summary-level data from large GWAS and Mendelian randomization analyses to examine the potential reciprocal relationships between AC levels and depression. We showed that genetically predicted lower levels of short-chain C2 (acetylcarnitine) and C3 (propionylcarnitine) and higher levels of medium-chain C8 (octanoylcarnitine) and C10 (decanoylcarnitine) were associated with increased depression risk. No reverse impact of depression liability on AC levels was found.

In the present study, we carried forward these four ACs and tested whether the relationships predicted from genomic data were expressed in actual phenotypes measured in a large cohort (N∼2500) with extensive clinical phenotyping. We examined the association of ACs blood levels with the presence of MDD and with overall depression severity. We also explored whether this association varied across different symptom profiles. Previous research has shown that immuno-metabolic dysregulations map more consistently to symptoms of the “atypical” spectrum characterized by altered energy intake/output (in particular the reversed neurovegetative symptoms of hyperphagia, hypersomnia with leaden paralysis and fatigue) ^[2]^ and symptoms of anhedonia ^[14]^. We examined the associations of AC levels with three symptom profile scores of atypical/energy-related, anhedonic and melancholic symptoms. Finally, to distill the most consistent and reliable associations, we replicated the analyses from the same subjects with the same measures collected at 6-year follow-up and pooled estimates (>4000 observations) obtained at the two assessment timepoints.

## METHODS

### Study design and Setting

Data were obtained from the Netherlands Study of Depression and Anxiety (NESDA), an ongoing naturalistic longitudinal cohort study examining course and consequences of depressive and anxiety disorders. A description of the study rationale, design, and methods is given elsewhere^[15]^. Briefly, in 2003-2007, 2981 participants were recruited from community settings, primary care practices and mental health care institutions and were followed-up during biennial assessments. During the 9-year follow-up (2014–2017), full-biological siblings of NESDA participants with a lifetime affective disorder were additionally recruited. Participants were excluded if they had a self-reported diagnosis of psychiatric disorders not subject of NESDA (e.g. bipolar, psychotic, or cognitive disorders) or were not fluent in Dutch. All participants provided written informed consent and the study was approved by the Medical Ethics Committees of all the participating universities.

From the 3348 subjects of the NESDA cohort, we selected healthy controls and those with a diagnoses of MDD at their baseline assessment and we excluded those with only diagnoses of dysthymic disorder (n=66) or anxiety disorder (n=508). Then, we excluded the subjects with missing data for the four investigated metabolites (n=334) and for the depressive symptom severity questionnaire (n=34). Thus, the main analytical sample included 2574 participants (2363 from NESDA baseline and 211 siblings) with data on MDD diagnostic status, overall depression severity, and depressive symptoms profiles and at least one of the investigated metabolites. Furthermore, additional data from 1621 subjects with the same measures of depression and metabolites repeated at 6-year follow-up (2010-2013) were used for internal replication.

### MDD diagnostic status, overall depression severity and profiles

The presence of DSM-IV diagnosis of MDD was assessed using the Composite Interview Diagnostic Instrument version 2.1^[16]^ administered by specially trained research staff. Three groups were identified: participants with current MDD (that is, within the past 6 months), with remitted (lifetime but not current) MDD, and healthy controls (without any lifetime depressive/anxiety disorder). Overall depression symptom severity was measured with the Inventory of Depressive Symptomatology self-report questionnaire (IDS-SR_30_^[17]^), with scores ranging from 0 to 84.

Three depression symptom profiles were created using items from the IDS-SR_30_ as described in previous studies^[2,18,19]^: 1) the atypical energy-related symptom (AES) profile based on the five items of hypersomnia, increased appetite, increased weight, low energy and leaden paralysis (range 0-15); 2) the anhedonic profile based on three items of response of mood to good or desired events, general interest and capacity for pleasure or enjoyment (range 0-9); and the melancholic profile based on eight items of waking up too early, quality of mood, hypophagia, decreased weight, linkage of mood to time of day (if worse in the morning), view of self, psychomotor agitation and psychomotor retardation (range 0-24).

### Metabolomics profiling and data processing

After an overnight fast, EDTA plasma samples were collected and stored in aliquots at -80°C until further analysis. Samples were sent in two shipments to the USA. Metabolic profiles were measured using the untargeted metabolomics platform from Metabolon Inc (Durham, NC). Extended description of the assessment is provided elsewhere^[20]^ and in Supplemental methods. Three of the four ACs (C2, C8 and C10) investigated in the present study were not available in the previously described^[20]^ metabolomics dataset. Issues with batch normalization using NIST samples as reference due to low levels of these ACs in NIST compared to NESDA samples led to exclusion of these measures according to our quality control criteria after batch correction. For the present analyses, we re-processed the raw measurements of these three ACs using their median ion counts in each batch for normalization. Applying this approach, coefficients of variation (CVs) of plasma reference samples that were run along with the NESDA samples were below 25% for the four measures, thereby meeting the original quality control criteria. Batch-normalized values of the four AC metabolites were log2-transformed and metabolite levels higher than 3[standard deviations (C2 1.30%; C3 0.82%; C8 0.90%; C10 0.70%) were set as missing.

### Covariates

Covariates included age, sex, educational level and metabolomic assessment shipment (first vs second). Health and lifestyle information included smoking status (non-smoker vs current smoker), alcohol consumption as units per week, physical activity assessed using the International Physical Activity Questionnaire (IPAQ)^[21]^ (expressed in Metabolic Equivalent Total (MET) minutes per week), and body mass index (BMI). The number of self-reported current somatic diseases (including cardiometabolic, respiratory, musculoskeletal, digestive, neurological, endocrine diseases and cancer) for which participants received medical treatment was counted (coded as 0, 1, 2+) as a global marker of poor physical health. In specific secondary analyses we examined the impact of antidepressant use, measured based on drug container inspection of medications used in the past month, classified according to the World Health Organization Anatomical Therapeutic Chemical classification: selective serotonin reuptake inhibitors (N06AB), tricyclic antidepressants (N06AA) and other less commonly prescribed medications (N06AX, N06AF, N06AG).

### Statistical methods

Variables were reported as percentages or means ±SD as appropriate. Pairwise correlation between depressive symptom scores were estimated with Pearson’s r coefficient.

All analyses were performed using linear mixed models with “family-factor” as random effect, in order to take into account the pedigree structure of the sample. We initially tested differences in adjusted mean AC levels across the three diagnostic groups: current MDD, remitted MDD and healthy controls. Adjusted mean AC levels across the three groups were estimated from the mixed models, standardized differences between groups were reported using Cohen’s *d* tested in post-hoc pairwise comparisons. To estimate the association between metabolites and overall depression severity, we regressed AC levels on IDS-SR_30_ total scores. Metabolite levels and depressive symptom scales were expressed as SD unit increase to derive standardized estimates. To further explore the functional shape of this association we applied restricted cubic splines with 3 knots to regression models. We examined the potential impact of antidepressant use on previous analyses by repeating the models excluding participants on antidepressants. ACs significantly linked to MDD status and/or depression severity were carried forward in subsequent analyses examining whether the association was mainly driven by specific symptom profiles, by regressing the metabolite levels on each of the three symptom profile scales.

All models were adjusted for age, sex, educational level, and shipment. For significant associations, we tested the potential explanatory effect of lifestyle and health-related variables by further including alcohol consumption, smoking status, physical activity, BMI and the number of self-reported current somatic diseases in the analytical models.

Lastly, to evaluate the consistency of the associations detected, we performed internal replication analyses using data from the 6-year follow-up followed by pooling of all observations (N=4195) in a unified mixed model with two random factors (one for the family effect and one for the repeated observations from the same subject over time).

Analyses were performed in *RStudio* version 2023.03.0+386 (RStudio: Integrated Development for R). All statistical tests were two-sided and used a significance level of *P*<0.05. In main analyses, False-Discovery Rate (FDR) q-values were calculated. The present study report follows the STROBE (Strengthening the Reporting of Observational Studies in Epidemiology) Statement (Supplementary Table S5).

## RESULTS

The sample’s mean age was 42.8 years (SD 13.19) and 65.3% were females (Table 1). Participants had current MDD (N=1035), remitted MDD (N=739) or were healthy controls (N=800); the mean IDS-SR_30_ score was 20.01±13.98. Distributions of metabolite levels are depicted in Supplementary Figure S1 and pairwise correlation between metabolites are shown in Supplementary Figure S2. C8 and C10 were highly correlated (Person’s r=0.9) in line with previously reported genetic correlations (rg=0.98)^[13]^ while all other pairs showed Person’s r <0.5.

**Table 1:** General characteristics of the sample at baseline (n=2574)

| **Table 1:** General characteristics of the sample at baseline (n=2574) |  |
| --- | --- |
| *Sociodemographic characteristics* |  |
| Age, years - mean±SD | 42.80±13.19 |
| Gender - n(%) |  |
| Male | 892 (34.7) |
| Female | 1682 (65.3) |
| Level of education, years - mean±SD | 12.19±3.27 |
| *Lifestyle and health* |  |
| Smoking status - n(%) |  |
| No smoker | 1636 (63.6) |
| Current smoker | 938 (36.4) |
| Physical activity - mean±SD |  |
| MET total | 3752.16±3097.32 |
| Chronic diseases - n(%) |  |
| None | 1174 (45.6) |
| One disease | 819 (31.8) |
| 2 or more | 581 (22.6) |
| Alcohol use, ml/week (mean±SD) | 7.04±9.66 |
| BMI - mean±SD | 25.53±4.77 |
| *Clinical characteristics* |  |
| Diagnostic status – n (%) |  |
| MDD current (past 6 months) | 1035 (40.2) |
| MDD remitted | 739 (28.7) |
| HC | 800 (31.1) |
| Antidepressant use, yes – n(%) | 642 (24.9) |
| Severity of MDD, IDS-SR<sub>30</sub> total score | 20.01±13.98 |
| SD, standard deviation; MET, Metabolic Equivalent of Task; BMI, Body Mass Index; MDD, Major Depressive Disorder; HC, Healthy Control; IDS-SR<sub>30</sub>, Inventory of Depressive Symptoms, Self Rated 30 items |  |

### Differences in ACs across MDD groups

Figure 1 shows the age-, sex-, education-, and shipment-adjusted means of the four metabolite levels across the three diagnostic groups. A significant overall difference across groups was found only for C2 levels (overall-p=1.05e-5, q=4.26e-5): current (mean=-0.06, SE=0.01) and remitted (mean=-0.05, SE=0.03) MDD cases had similar significantly lower mean C2 levels (*d*=-0.2) as compared to controls (mean=0.03, SE=0.06). This difference remained statistically significant after further adjustment for alcohol consumption, smoking status, physical activity, BMI and number of somatic diseases (current MDD mean=-0.06, SE=0.01; remitted MDD mean=-0.06, SE=0.02; healthy controls mean 0.03, SE=0.02; overall-p=3.14e-5, q=1.26e-4). Of interest, C3 levels were increasingly elevated from healthy controls (mean=0.63, SE=0.02) to subjects with remitted (mean=0.65, SE= 0.02) to those with current (mean=0.68, SE=0.02) MDD, although the overall difference was not statistically significant (overall-p=1.65e-1, q=2.32e-1).

**Figure 1.**
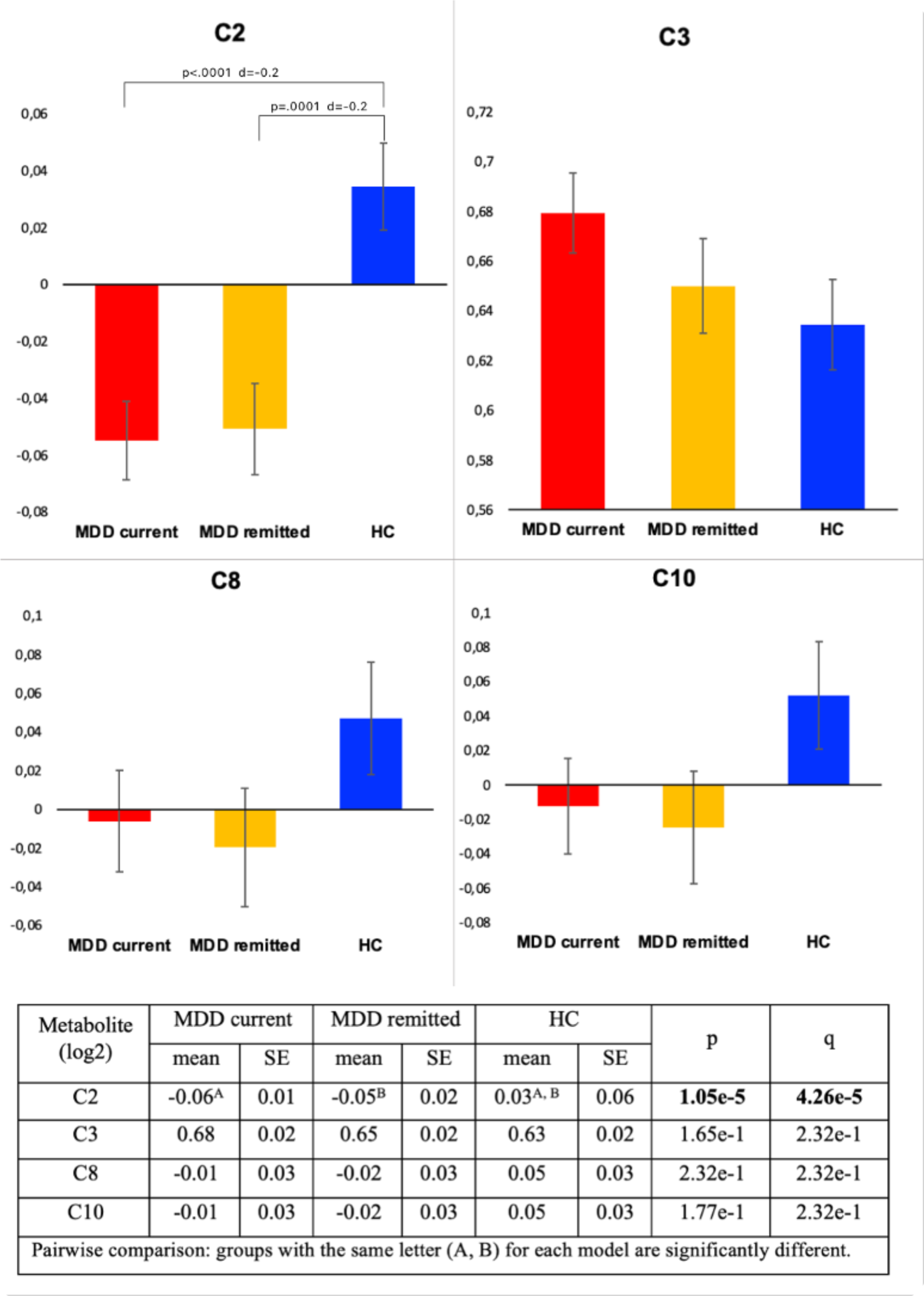
Metabolite levels across the three groups: current MDD (n=1035), remitted MDD (n=739) and healthy controls (n=800). Y-axes: metabolite log2 levels. Analysis were adjusted for age, sex, education and shipment. Adjusted P-levels (q) are obtained by BH-FDR (Benjamini-Hochberg False Discovery Rate) correction, used to control the rate of false positives in multiple testing.

Participants using antidepressants were 24.9% of the total sample, and specifically 44.9% of those with current MDD and 31.5% of those in the remitted group. To evaluate whether the identified association between C2 and MDD status was driven by antidepressant use, we repeated the analysis only considering participants who did not use antidepressants (Table S1). The differences in C2 levels between cases (both current and remitted) versus healthy controls was confirmed to be similar (d=-0.2) and statistically significant (overall-p=1.20e-3).

### Association of ACs with overall depression severity

We estimated the association between AC levels and overall severity of depression measured by IDS-SR_30_ total score (Table S2). Higher depression severity was significantly associated only with higher C3 levels (ß=0.06, SE=0.02, p=1.21e-3, q=4.80e-3) correcting for age, sex, education and shipment. Although slightly reduced, the association was still statistically significant (ß=0.04, SE=0.02, p=2.67e-3) in the fully adjusted model. To examine the potential impact of antidepressant use, we re-estimated the association between C3 and depression severity in participants without such medications (N=1916): results were substantially similar (ß=0.06, SE=0.02, p=1.16e-2). In line with previous analyses examining MDD diagnostic status, we found a negative association between overall depressive severity and C2 levels, although not reaching statistical significance (ß=-0.03, SE=0.02, p=8.10e-2, q=1.62e-1). Furthermore, IDS-SR_30_ scores were not significantly associated with C8 (ß=-0.001, SE=0.02, p=9.48e-1, q=9.48e-1) or C10 (ß=-0.002, SE=0.02, p=9.26e-1, q=9.48e-1).

To study the apparently discordant results for C2 and C3 in analyses using MDD diagnostic status versus those with continuous symptom severity, we examined the functional shape of the association between depression severity and ACs by fitting restricted cubic splines (3 knots) regression models. As shown in Figure 2A, the fitted spline (adjusted for age, sex, education and shipment) revealed a threshold in the symptoms-C2 relationship, becoming inversely associated mainly for IDS-SR_30_ scores values below the sample mean. Healthy controls had mean IDS-SR_30_ of 1 SD above the mean (the threshold point of the association), while MDD cases with remitted and those with current MDD had mean value, respectively, around the mean and 1 SD below it. This shows how this non-fully-linear relationship was better captured by the analyses employing categorical diagnostic groups. In contrast, the association between IDS-SR_30_ scores and C3 appeared substantially linear (Figure 2B), thus potentially better captured in analyses using continuous depressive symptom scores.

**Figure 2.**
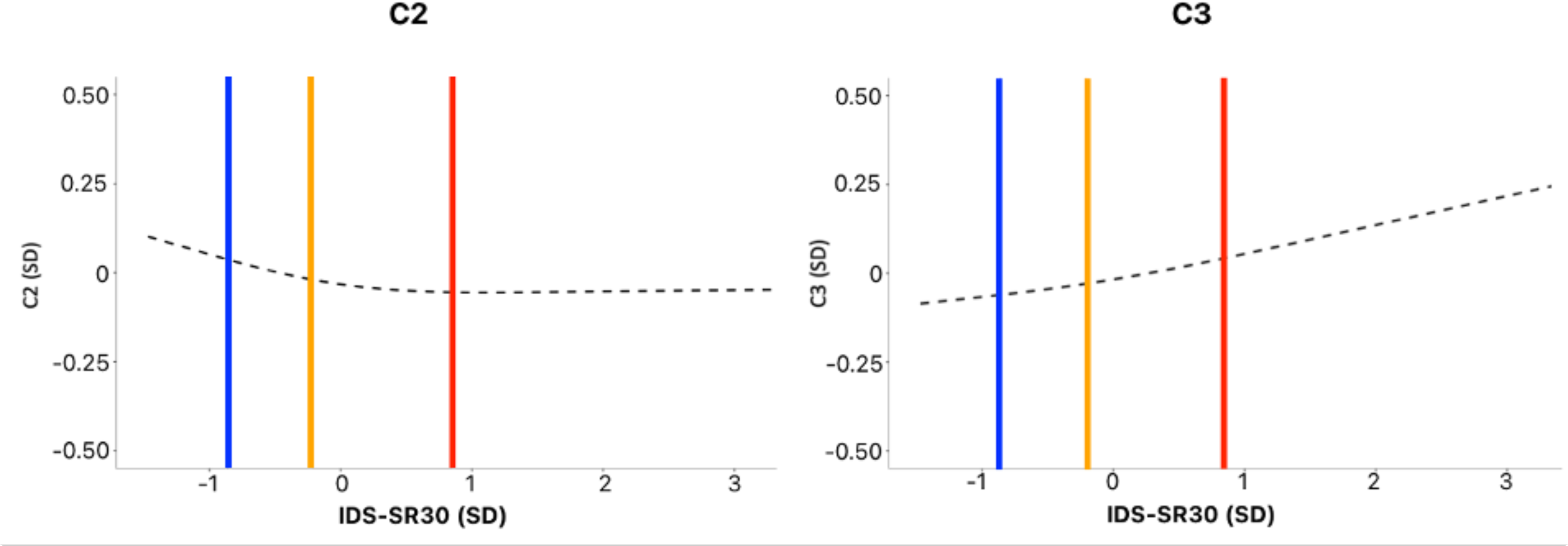
Restricted cubic spline for C2 and C3. Dashed fitted spline of C2 across IDS-SR_30_ scores. Vertical lines illustrate the mean IDS-SR_30_ values across the three diagnostic groups: current MDD (red line), remitted MDD (yellow line), HC (blue line).

### Associations of ACs with symptom profiles

We further evaluated whether the identified associations were specific for certain symptom profiles. There were moderate positive correlations between the three symptom profiles (Figure S3) varying from r=0.75 (anhedonic and melancholic profiles) to r=0.56 (AES and melancholic profiles), indicating that the scales captured partially non-overlapping dimensions.

The AES profile score was significantly associated with lower C2 (ß=-0.05, SE=0.02, p=1.85e-2, q=3.70e-2) and higher C3 (ß=0.08, SE=0.02, p=3.41e-5, q=2.05e-4) levels (Table 2). These associations remained statistically significant after further adjustment for lifestyle and health-related variables (C2 ß=-0.04, SE=0.02, p=3.92e-2; C3 ß=0.04, SE=0.02, p=3.13e-2).

**Table 2.** Association of metabolites levels with diagnostic status, MDD severity and symptom profiles. Results for the main analysis (n=2574), internal replication (n=1567) and total pooling (n=4141) are reported.

| Metabolite |  |  | Main analyses<br>(n=2574) |  |  | Internal replication<br>(n=1621) |  |  | Total pooling<br>(n=4195) |  |  |
| --- | --- | --- | --- | --- | --- | --- | --- | --- | --- | --- | --- |
| | | | $\beta$ | SE | P | $\beta$ | SE | P | $\beta$ | SE | P |
| C2 | Diagnostic status | Current MDD | -0.21 | 0.05 | **1.16e-5** | -0.01 | 0.07 | 8.91e-1 | -0.15 | 0.04 | **4.92e-4** |
|  |  | Remitted MDD | -0.20 | 0.05 | **8.67e-5** | 0.05 | 0.05 | 3.51e-1 | -0.13 | 0.04 | **1.57e-3** |
|  | MDD severity | IDS-SR<sub>30</sub> total | -0.03 | 0.02 | 8.10e-2 | 0.03 | 0.02 | 1.72e-1 | -0.02 | 0.02 | 2.56e-1 |
|  | Symptom profile | AES | -0.05 | 0.02 | **1.85e-2** | 0.01 | 0.02 | 7.59e-1 | -0.03 | 0.02 | **3.45e-2** |
|  |  | Anhedonic | -0.01 | 0.02 | 4.98e-1 | 0.03 | 0.02 | 2.15e-1 | -0.002 | 0.02 | 8.37e-1 |
|  |  | Melancholic | -0.01 | 0.02 | 4.76e-1 | 0.04 | 0.02 | 1.12e-1 | -0.002 | 0.02 | 9.63e-1 |
| C3 | Diagnostic status | Current MDD | 0.08 | 0.04 | 6.01e-2 | 0.003 | 0.05 | 9.57e-1 | 0.07 | 0.04 | 1.37e-1 |
|  |  | Remitted MDD | 0.03 | 0.05 | 5.42e-1 | 0.04 | 0.04 | 3.67e-1 | 0.07 | 0.04 | 2.01e-1 |
|  | MDD severity | IDS-SR<sub>30</sub> total | 0.06 | 0.02 | **1.21e-3** | 0.03 | 0.02 | 1.15e-1 | 0.04 | 0.02 | **1.85e-2** |
|  | Symptom profile | AES | 0.08 | 0.02 | **3.41e-5** | 0.05 | 0.02 | **4.37e-3** | 0.04 | 0.01 | **2.32e-3** |
|  |  | Anhedonic | 0.05 | 0.02 | **5.75e-3** | 0.004 | 0.02 | 8.13e-1 | 0.02 | 0.01 | 9.57e-2 |
|  |  | Melancholic | 0.03 | 0.02 | 7.67e-2 | 0.01 | 0.02 | 4.01e-1 | 0.004 | 0.01 | 7.68e-1 |

Covariates – shipment, age, sex, education.
n, number of observations; SE, Standard error; P, level of significance; AES, atypical energy-related symptoms.

The anhedonic profile score was associated with higher C3 (ß=0.05, SE=0.02, p=5.75e-3, q=1.73e-2) levels, although the association was reduced (ß=0.03, SE=0.02, p=5.98e-2) after additional adjustment for lifestyle and health-related variables. The melancholic profile score was not significantly associated with C2 or C3. No associations were found for C8 and C10 (Table S3).

To consider the possible non-linear shape of the relationship between C2 levels and symptom profile scales, we performed additional analysis for C2 dividing the profile scores in tertiles. Analyses confirmed the association between C2 and AES (Table S4) and its non-fully linear functional form (Figure S4).

### Internal replication and pooled analyses

Significant associations detected using baseline data were further tested with an internal replication sample of 1621 subjects with metabolites and depressive symptoms assessed at the 6-year follow-up. At this later wave of assessment, participants had an improved clinical profile as compared to baseline, with a lower proportion of current MDD (16% versus 40%) diagnoses and lower IDS-SR_30_scores (15.21±12.09 versus 20.0±13.98). In the internal replication sample, estimates were consistent with those in the main analyses, but only the association between the AES profile score and C3 was statistically significant (ß=0.05, SE=0.02, p=4.37e-3). The overall model pooling 4195 observations from main and internal replication samples (Table 2) confirmed the associations between C2 and current MDD (ß=-0.15, SE=0.04, p=4.92e-4), remitted MDD (ß=-0.13, SE=0.04, p= 1.57e-3) and with the AES profile (ß=-0.03, SE=0.02, p=3.45e-2). For C3, the model confirmed the association with overall (ß=0.04, SE=0.02, p=1.85e-2) and AES (ß=0.04, SE=0.01, p=2.32e-3) symptom scores.

## DISCUSSION

This is the largest study to date to explore the relationship between ACs blood concentrations and depression in a cohort enriched of clinical cases psychiatrically well characterized. Alterations of small effect size in levels of short-chain ACs, reduced acetylcarnitine (C2) and elevated propionylcarnitine (C3), were linked to the presence and intensity of depression. Results were confirmed in pooled analyses with >4,000 observations additionally including an internal replication sample with data collected at 6-year follow-up.

Findings from a previous study^[13]^ leveraging genomic data and Mendelian randomization analyses suggested a potential causal relationship between low C2 and depression risk. Consistently, in the present study lower C2 levels as compared to healthy controls were observed for subjects experiencing a current depressive episode as well those who had remitted. This pattern supports the interpretation of decreased C2 levels as a potential “trait marker” indexing an underlying vulnerability for the development of depression, in line with previous genetic analyses^[13]^. Nevertheless, lower C2 in remitted subjects could also represent the result of a “scar effect” of depression, not improving after symptomatologic remission. Further longitudinal analyses in initially non-depressed subjects are needed to properly disentangle these two mutually non-exclusive scenarios.

Furthermore, we found that C3 levels were positively correlated with the severity of depression, acting as a potential “state marker” of the current symptomatology. This finding is in contrast with the expectation from Mendelian randomization analyses, showing an association between genetically predicted lower C3 levels and depression risk. Discrepancies between analyses employing genetic instruments and actual phenotypes may provide intriguing insights on relevant dynamics. For instance, it could be speculated that such discrepancies may reflect compensatory mechanisms aimed at correcting underlying vulnerability, consistently with previous findings ^[22]^ showing an increase in C3 during antidepressant treatment. Presently such hypothesis remains merely speculative; to reconcile the genetics and observational estimates additional longitudinal and experimental studies are needed. Intriguingly, the direction of the association with depression was opposite for the two short-chain ACs C2 and C3. Although having a partially overlapping genetic basis, the actual correlation of C2 and C3 blood concentrations was weak (r=0.2) and these ACs are components of partially independent pathways (Figure 3). C2 is a downstream product of mitochondrial beta-oxidation of long-chain fatty acids. Disruptions in this process may result in reduced in C2 levels. C3 is a downstream product of the metabolism of branched-chain amino-acids (leucine, isoleucine, valine) and of odd-chain fatty acids. In diseases where amino-acid metabolizing enzymes are dysfunctional or absent (e.g., propionic acidemia and methylmalonic acidemias characterized by neurological symptoms, muscle weakness and low energy) C3 accumulates and higher blood levels are used as a screening tool. Hence, it is possible that C2 and C3 play distinct roles in various molecular pathways associated with depression. Further research is required to gain a more comprehensive understanding of this aspect.

**Figure 3.**
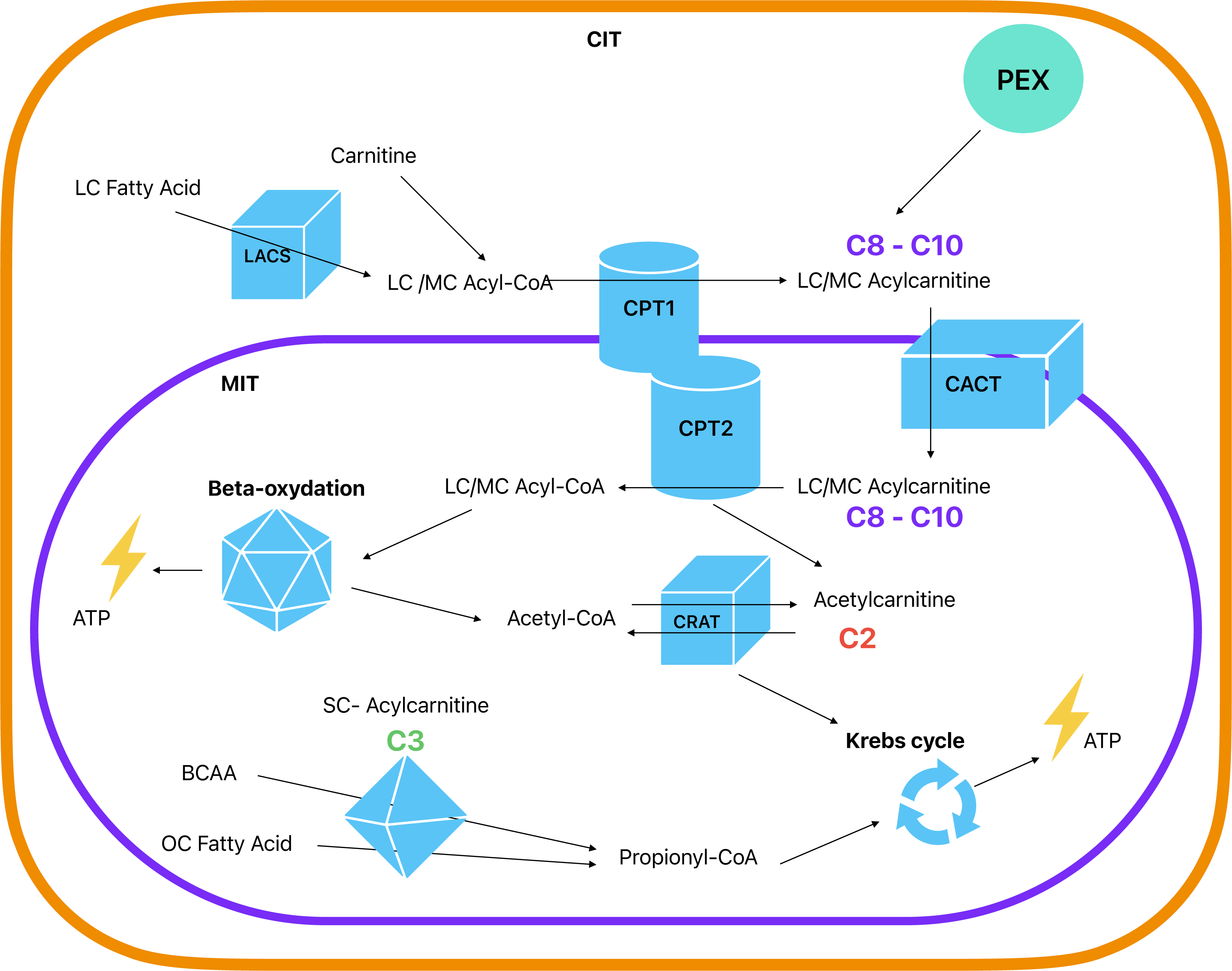
Acylcarnitine production and main roles in mitochondrial function. Abbreviations: CIT, cytosol; MIT, mitochondria; PEX, peroxisome; LC, long-chain; MC, medium-chain; SC, short-chain; CoA, coenzyme A; LACS, long-chain acyl-coenzyme A synthetase; CACT, carnitine/acylcarnitine translocase; CPT1 and CPT2, carnitine palmitoyl-transferase 1 and 2; CRAT, carnitine O-acetyltransferase; ATP, adenosine triphosphate; BCAA, branched-chain amino acid; OC Fatty Acid, Odd-chain fatty acid. Model based on Dambrova et al. (2022)^[44]^ and Li et al. (2019)^[45]^. Full description in online supplement.

Finally, while genetically predicted higher levels of the medium-chain ACs C8 and C10 were associated with increased depression risk in previous Mendelian randomization analyses, no significant association with depression presence or severity was found for levels of these ACs in the present study. Potential compensatory mechanisms correcting underlying vulnerability may be speculated as one of the reasons for such discrepancies, consistently with previous findings showing decrease in C8 and C10 after antidepressant treatment ^[22]^ Nevertheless, other conceptual and methodological differences may explain discrepancies in results between genetically informed (capturing average lifetime risk and etiological mechanisms) and observational (capturing time specific or acute events and disease progression) analyses. As addressed for other mental disorders, the integration of static genetic data and dynamic “omics” data is necessary to define better biomarkers for clinical management^[23]^. Furthermore, the network of biological pathways involving ACs and converging on the mitochondria is extremely complex (Figure 3). Rather than alterations in single components, the impact on depression pathobiology may be due to the net effect of different dysregulations and interrelated compensatory mechanisms in such a complex network, which could be fully unraveled only by further functional and mechanistic studies.

The present findings are in line with previous evidence suggesting that metabolic alterations are not uniformly associated with all clinical manifestation of depression, but map more consistently with specific symptom profiles^[24]^. Across different analyses, lower C2 and higher C3 levels were associated with an atypical/energy-related symptom profile characterized by altered energy intake/expenditure balance (e.g. hyperphagia, weight gain, hypersomnia, fatigue, leaden paralysis) and previously shown to be linked^[2,18,25]^ to inflammatory and metabolic alterations. Less consistent evidence across analyses were found for an association between higher C3 levels and an anhedonic symptom profile previously linked with inflammation and neurobiological reward processes. The clustering between specific biological and clinical features has been postulated to identify a theoretical dimension labelled “immuno-metabolic depression (IMD)”^[2]^ (aligning with the Research Domain Criteria (RDoC) framework^[26]^) that may be conceptualized as a depression dimension in mapping the degree of expression of transdiagnostic bio-behavioral processes overlapping with those of other constructs (e.g. sickness behavior) ^[27,28]^, psychiatric diagnoses (e.g. bipolar disorder, seasonal affective disorder) or somatic (e.g. cardiovascular diseases, diabetes) conditions. In this context, engagement of the specific immuno-metabolic biological pathways (e.g. AC metabolism) in conjunction with the expression of specific clinical symptoms (e.g. atypical/energy-related, anhedonia) may identify depressed subjects at higher cardiometabolic risk. For example, altered levels of short-chain ACs, including C2 and C3, have been observed in coronary artery disease and diabetes^[6,29]^.

Findings from the present study align with hypotheses^[3]^ suggesting that mitochondrial energetic dysfunction may be involved in the pathophysiology of depression. Cellular energy dysfunction may contribute to depression through various pathways, leading to neurotoxicity and impaired neuroplasticity^[4,30]^. In animal models, C2 supplementation promoted neuroplasticity, synthesis of neurotrophic factors, modulation of glutamatergic dysfunction, reversal of neuronal atrophy in regions like the hippocampus and amygdala, and improvement in depression-like behavioral symptoms^[31–34]^.

Moreover, mitochondrial dysfunction and related oxidative stress can activate the innate branch of the immune system, leading to the release of pro-inflammatory cytokines influencing various depression-related pathophysiological mechanisms: monoaminergic neurotransmission disruption, tryptophan degradation toward neurotoxic catabolites, glutamate-related excitotoxicity, decreased neurotrophic factors and alterations in the hypothalamic-pituitary-adrenal axis^[3,14,35]^. Mitochondrial bioenergetic dysfunctions may also have broader impact on immune processes. It has been recently shown^[36]^ that T cells from depressed patients displayed a compromised metabolic profile accompanied by heightened gene expression of CTP1a (carnitine palmitoyltransferase 1A), the mitochondrial enzyme responsible for ACs synthesis.

It is important to acknowledge that the relationships we found between AC levels, MDD diagnosis, depression severity and depression profiles may be explained to some extent by shared distal environmental and lifestyle factors (e.g., comorbid somatic diseases, sedentary behavior, smoking, high-fat diet, alcohol consumption^[37–40]^) that could act as confounder or mediators of the association. Nevertheless, when analyses were adjusted for BMI, level of physical activity, number of somatic comorbidities, smoking status and alcohol use, results were substantially unchanged.

Alternatively, altered levels of ACs may be a direct consequence of depression or related clinical aspect such as use of antidepressants. Nevertheless, previous genetic study employing Mendelian randomization^[13]^ found no evidence supporting a causal role of depression liability in influencing AC levels. Furthermore, in the present study we repeated the analysis excluding subjects using antidepressants and associations were not substantially impacted.

A major limitation of the present study is the cross-sectional design of the analyses, which estimated the associations between AC levels and depression at the same assessment (either baseline or 6-year follow-up), precluding conclusions about causality. Strengths of the current study are the large sample size and the detailed clinical assessment of depression and related characteristics. A unique strength is represented by the internal replication sample with data collected from the same subject at a different time point (>4,000 observations), supporting the consistency and reliability of the associations detected. Nevertheless, the present findings warrant external replication in independent samples with similar measures when these become available. At the same time, it is important to remark that the finding of lower C2 as risk factor for depression is highly consistent with data from other studies using clinical samples, animal models and genomic data^[11,13]^.

In the future, longitudinal studies will be necessary to capture trajectories of changes over time in AC levels and depressive symptoms in order to properly disentangle trait vs state effects and provide empirical grounding for causal interpretation, triangulating evidence with experimental medicine approaches. To date, only few small studies with heterogeneous methodology tested carnitine/acetylcartinitine supplementation in depressed patients, producing inconsistent results^[41]^. In parallel, in-depth mechanistic studies could identify the precise biological mechanisms underlying the association between ACs and depression. An interesting approach would be to study differences in ratios between long, medium and short chain ACs in order to assess possible alterations in enzymatic function in depressed patients, a method already used in other fields of medicine^[42,43]^.

In conclusion, the present study identified alterations of small effect size in blood levels of short-chain ACs related to the presence and severity of depression, especially of clinical profiles expressing symptoms reflecting altered energy homeostasis. Cellular metabolic dysfunctions may represent the biological substrate connecting depression with different cardiometabolic outcomes and a key pathway in depression pathophysiology potentially accessible through AC metabolism.

## Data Availability

The data that support the findings of this study are available on request from the NESDA cohort study. The data sharing policy of NESDA and information on how to request for access to study data is available at https://www.nesda.nl/nesda-english/.

## Acknowledgments

The infrastructure for the NESDA study (http://www.nesda.nl) is funded through the Geestkracht program of the Netherlands Organisation for Health Research and Development (ZonMw, grant number 10-000-1002) and financial contributions by participating universities and mental health care organizations (VU University Medical Center, GGZ inGeest, Leiden University Medical Center, Leiden University, GGZ Rivierduinen, University Medical Center Groningen, University of Groningen, Lentis, GGZ Friesland, GGZ Drenthe, Rob Giel Onderzoekscentrum).

R. Kaddurah-Daouk at Duke is PI of the Mood disorder Precision Medicine Consortium (funded by NIMH R01MH108348) and the Alzheimer Gut Microbiome Project (funded by NIA U19AG063744). She also received additional funding from NIA that enabled her research (RF1AG058942, RF1AG059093, U01AG061359, and R01AG081322).

Y. Milaneschi received funding from Amsterdam UMC (Starter Grant 2023).

M. Arnold and G. Kastenmüller received funding (through their institutions) from the National Institutes of Health/National Institute on Aging through grants RF1AG058942, RF1AG059093, U01AG061359, U19AG063744, R01AG069901, and R01AG081322.

