## Supplemental materials for "Acylcarnitines metabolism in depression: association with diagnostic status, depression severity and symptom profile in the NESDA cohort"

***ONLINE SUPPLEMENT***

SUPPLEMENTARY METHODS	1

Metabolite profiling and data processing	2

SUPPLEMENTARY RESULTS	3

Supplementary Figure 1 (Figure S1). Distribution of metabolite levels.	3

Supplementary Figure 2 (Figure S2). Levels of correlation of ACs metabolites.	4

Supplementary Figure 3 (Figure S3). Levels of correlation of the three symptom scales: atypical energy-related (AES), anhedonic and melancholic.	5

Supplementary Figure 4 (Figure S4). Restricted cubic spline for C2 and the three symptom profiles.	6

Supplementary Table 1. Estimated means (and SE) of C2 levels after excluding participants with antidepressant use.	7

Supplementary Table 2. Metabolite levels association with IDS-SR_30_ total scores.	8

Supplementary Table 3. Association of metabolite levels with symptom profile.	9

Supplementary Table 4. Association of C2 levels (divided in tertiles) with symptom profile.	10

Supplementary Table 5. STROBE Statement.	11

SUPPLEMENTARY DISCUSSION	13

Figure 3. Acylcarnitine production and main roles in mitochondrial function.	13

REFERENCES	14

### **SUPPLEMENTARY METHODS**

#### **Metabolite profiling and data processing**

After an overnight fast, EDTA plasma samples were collected and stored in aliquots at -80°C until further analysis. Samples were sent in two batches to the USA. Metabolic profiles were measured using the untargeted metabolomics platform from Metabolon Inc (Durham, NC)^[1]^. Briefly, plasma samples were divided into four fractions; two for ultra-high performance liquid chromatographytandem mass spectrometry (UPLC-MS/MS; positive ionization), one for UPLC-MS/MS (negative ionization), and one for a UPLC-MS/MS polar platform (negative ionization). Compounds were identified using an internal spectral database.

Peaks were quantified using the area-under-the-curve in the spectra as well as the raw metabolite data set, which was measured in 29 batches, included measures for 1367 metabolites in 5181 NESDA and 1008 reference samples (well characterized pooled human plasma samples: NIST SRM 1950 (n=288; 8-16 per batch) and Metabolon reference pooled plasma samples (n=720; 20-40 per batch)) and 1367 metabolites. NIST samples were used to calculate and control for technical measurement variability across the 29 batches. One experimental sample and one reference sample with high missingness (> 5SD + mean missingness) were excluded from the dataset. If outliers or apparent measurement issues within one plate or several plates within one batch were observed, all values on that plate were set to ‘NA’ to not affect subsequent batch correction. We batch corrected data by normalizing each metabolite value to the batch median of the metabolite measured in the NIST reference samples and then excluded those metabolites that still had a technical measurement variability of >30% using the Metabolon reference plasma samples. For metabolites, for which the metabolite was not detected in NIST samples, we used the overall batch median for normalization instead (affected metabolites are tagged). Next, we restricted the dataset to metabolites with missingness below 30% to ensure robust imputation results^[2]^. We then tested if missingness in any of the remaining 820 metabolites accumulated in one of the three measurement waves (baseline, 6-, or 9-year follow up) using a Fisher’s exact test. As this was not the case, we jointly imputed all waves using a k-nearest neighbor approach (k=10). Before statistical analysis, we log2 transformed the final dataset.

### **SUPPLEMENTARY RESULTS**

#### **Supplementary Figure 1 (Figure S1). Distribution of metabolite levels.**

Y-axes: frequency of metabolite level in total sample.

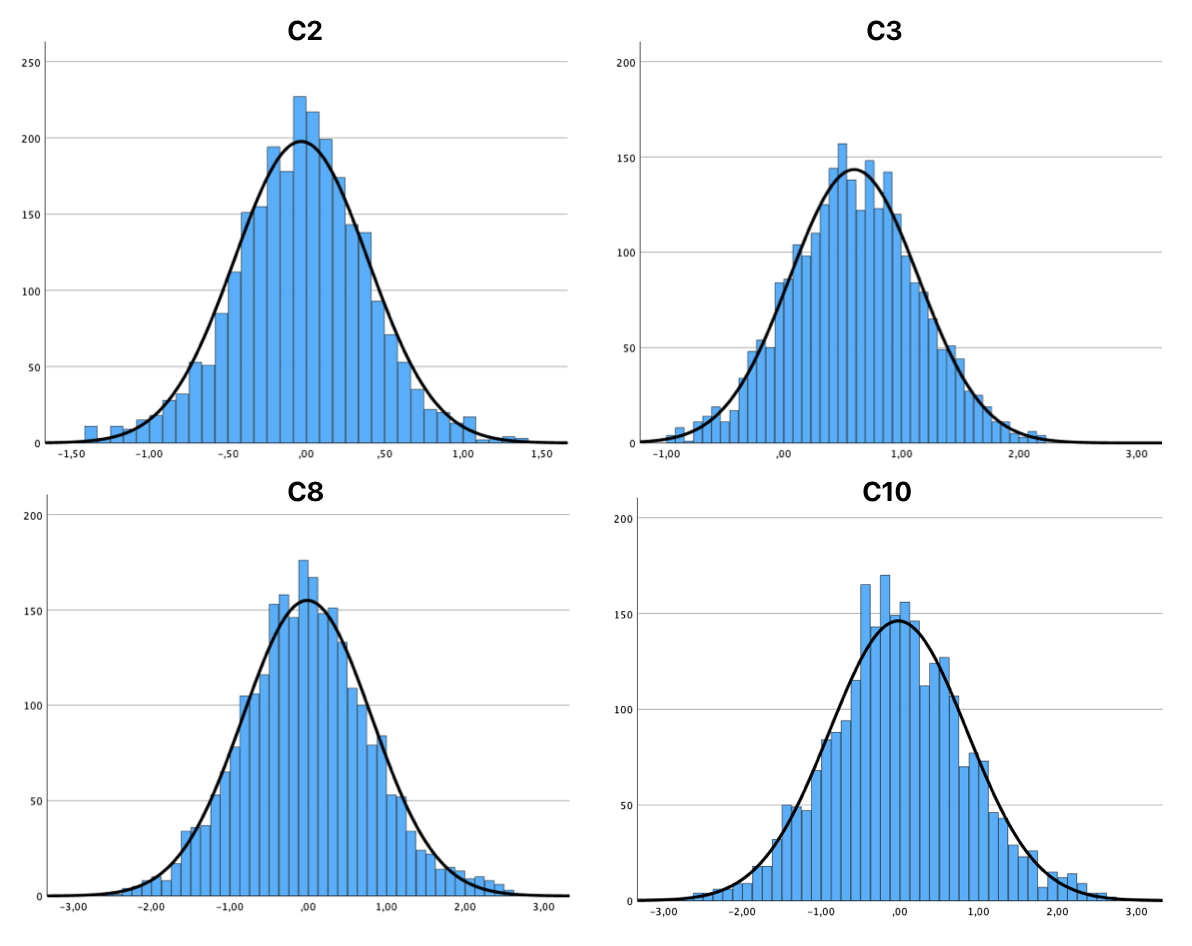

#### **Supplementary Figure 2 (Figure S2). Levels of correlation of ACs metabolites.**

The heatmap shows Person’s r correlation coefficients for each pair, with colour filling underlying the level of correlation, from 1 (red) to -1 (blue). All the correlations are statistically significant, with p<0.001.

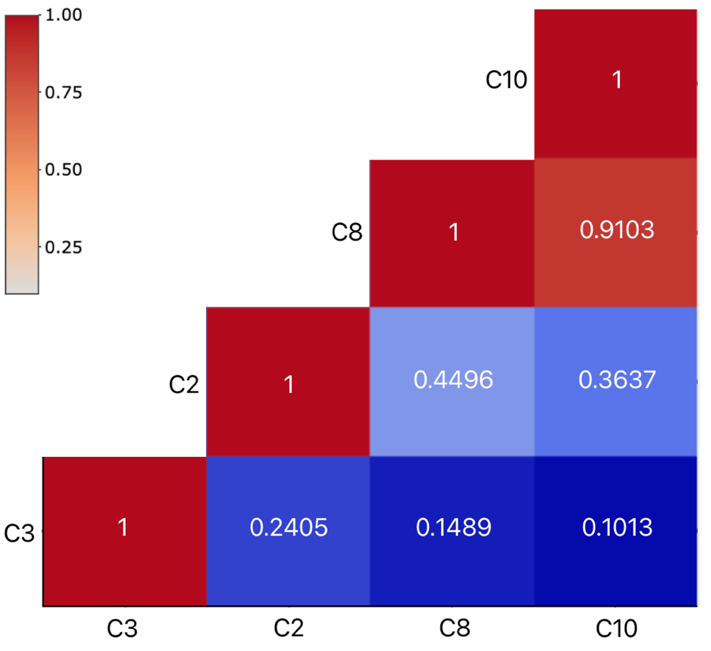

#### **Supplementary Figure 3 (Figure S3). Levels of correlation of the three symptom scales: atypical energy-related (AES), anhedonic and melancholic.**

The heatmap shows Person’s r correlation coefficients for each pair, with colour filling underlying the level of correlation, from 1 (red) to -1 (blue). All the correlations are statistically significant.

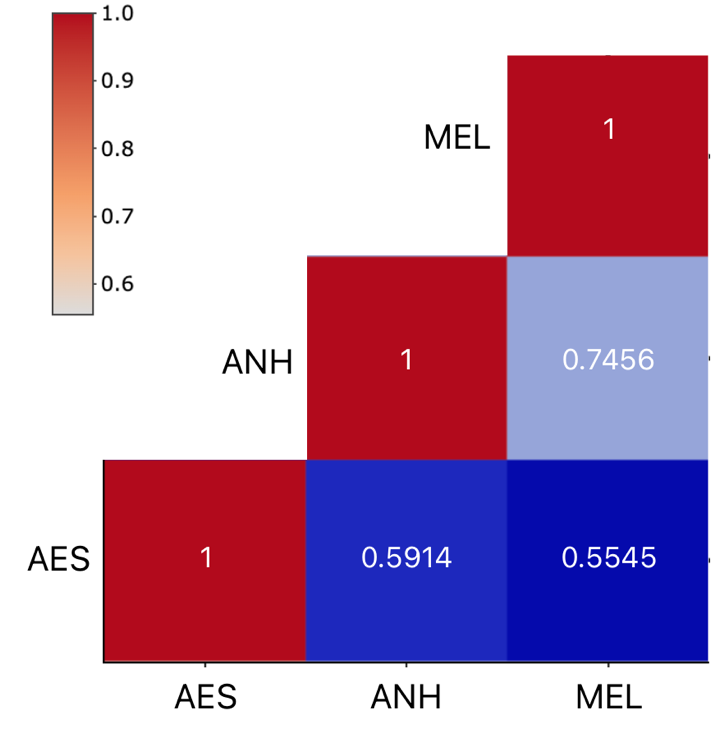

**Supplementary Figure 4 (Figure S4). Restricted cubic spline for C2 and the three symptom profiles.**

Dashed lines the fitted spline of C2 distribution across the three scales scores. Vertical lines illustrate the cutoff values that divide the scores in tertiles. AES: cut-offs -0.80 / 0.31; N I tertile 863 (34%), N II tertile 974 (38%), N III tertile 764 (30%). Anhedonic: cut-offs -0.54 / 0.28; N I tertile 1208 (47%), N II tertile 688 (27%), N III tertile 678 (26%). Melancholic: cut-offs -0.60 / 0.45; N I tertile 1057 (41%), N II tertile 760 (30%), N III tertile 757 (29%).

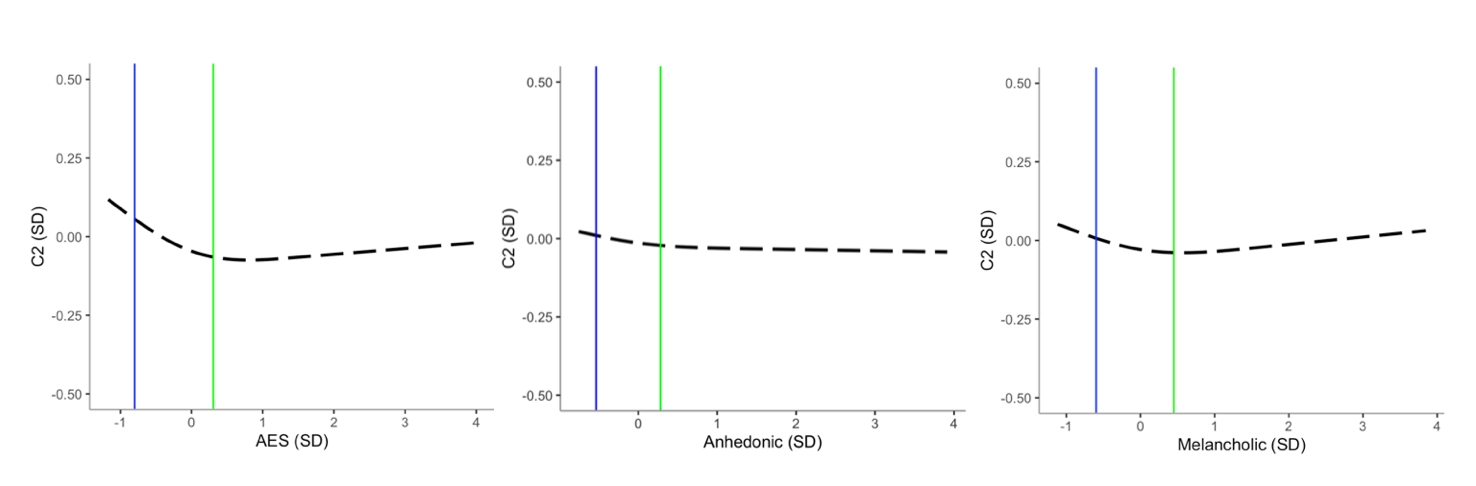

#### **Supplementary Table 1. Estimated means (and SE) of C2 levels after excluding participants with antidepressant use.**

| **Table S2.** Estimated means (and SE) of C2 levels among the three diagnostic status groups after excluding participants with antidepressant use (N=1916) | | | | | | | |
| --- | --- | --- | --- | --- | --- | --- | --- |
| Metabolite (log2) | MDD current | | MDD remitted | | HC | | P |
|  | mean | SE | mean | SE | mean | SE |  |
| C2 | -0.04^A^ | 0.02 | -0.03^B^ | 0.02 | 0.04^A,B^ | 0.02 | **1.20e-3** |
| Covariates –shipment, age, sex, education. | | | | | | | |
| Pairwise comparison: groups with the same letter (A, B) for each model are significantly different (A, P=9.0e-4, d=-0.2; B, P=5.3e-3, d=-0.2) | | | | | | | |
| MDD, major depressive disorder; HC, healthy control; P, level of significance; SE, standard error. | | | | | | | |

#### **Supplementary Table 2. Metabolite levels association with IDS-SR_30_ total scores.**

| **Table S3.** Metabolite levels association with IDS-SR_30_ total scores. | | | | |
| --- | --- | --- | --- | --- |
| Metabolite | Estimate | SE | P | q |
| C2 | -0.03 | 0.02 | 8.10e-2 | 1.62e-1 |
| C3 | 0.06 | 0.02 | **1.20e-3** | **4.80e-3** |
| C8 | -0.001 | 0.02 | 9.48e-1 | 9.48e-1 |
| C10 | -0.002 | 0.02 | 9.26e-1 | 9.48e-1 |
| Covariates – shipment, age, sex, education. SE, Standard error; P, level of significance; q, Adjusted P-levels obtained by BH-FDR (Benjamini-Hochberg False Discovery Rate) correction, used to control the rate of false positives in multiple testing. | | | | |

#### **Supplementary Table 3. Association of metabolite levels with symptom profile.**

| **Table S4.** Association of metabolite levels with symptom profile. | | | | | |
| --- | --- | --- | --- | --- | --- |
| Metabolite | Symptom profile | Estimate | SE | P | q |
| C2 | AES | -0.05 | 0.02 | **1.85e-2** | 7.40e-2 |
|  | Anhedonic | -0.01 | 0.02 | 4.99e-1 | 6.65e-1 |
|  | Melancholic | -0.01 | 0.02 | 4.77e-1 | 6.65e-1 |
| C3 | AES | 0.08 | 0.02 | **3.42e-5** | **4.14e-4** |
|  | Anhedonic | 0.05 | 0.02 | **5.80e-3** | **3.48e-2** |
|  | Melancholic | 0.03 | 0.02 | 7.67e-2 | 2.30e-1 |
| C8 | AES | -0.02 | 0.02 | 2.33e-1 | 4.66e-1 |
|  | Anhedonic | -0.002 | 0.02 | 9.35e-1 | 9.35e-1 |
|  | Melancholic | 0.01 | 0.02 | 5.89e-1 | 7.07e-1 |
| C10 | AES | -0.03 | 0.02 | 1.30e-1 | 3.12e-1 |
|  | Anhedonic | 0.01 | 0.02 | 7.40e-1 | 8.08e-1 |
|  | Melancholic | 0.02 | 0.02 | 3.00e-1 | 5.14e-1 |
| Covariates – shipment, age, sex, education. SE, Standard error; P, level of significance; AES, atypical energy-related symptoms; q, Adjusted P-levels obtained by BH-FDR (Benjamini-Hochberg False Discovery Rate) correction, used to control the rate of false positives in multiple testing. | | | | | |

#### **Supplementary Table 4. Association of C2 levels (divided in tertiles) with symptom profile.**

| **Table S5.** Association of C2 levels (divided in tertiles) with symptom profile.  AES: cut-offs -0.80 / 0.31; N I tertile 863 (34%), N II tertile 974 (38%), N III tertile 764 (30%).  Anhedonic: cut-offs -0.54 / 0.28; N I tertile 1208 (47%), N II tertile 688 (27%), N III tertile 678 (26%).  Melancholic: cut-offs -0.60 / 0.45; N I tertile 1057 (41%), N II tertile 760 (30%), N III tertile 757 (29%). | | | | | |
| --- | --- | --- | --- | --- | --- |
| Metabolite | Symptom profile | Tertiles | Estimate | SE | P |
| C2 | AES | I | -0.01 | 0.05 | **2.18e-2** |
|  |  | II | -0.01 | 0.05 | **3.28e-2** |
|  | Anhedonic | I | 0.01 | 0.05 | 8.55e-1 |
|  |  | II | -0.03 | 0.05 | 5.99e-1 |
|  | Melancholic | I | -0.05 | 0.05 | 3.21e-1 |
|  |  | II | -0.07 | 0.05 | 1.55e-1 |
| Covariates – shipment, age, sex, education. SE, Standard error; P, level of significance; AES, atypical energy-related symptoms | | | | | |

#### **Supplementary Table 5. STROBE Statement.**

| Table S6. STROBE statement. | | | |
| --- | --- | --- | --- |
|  | Item No. | Recommendation | Page  No. |
| **Title and abstract** | 1 | (*a*) Indicate the study’s design with a commonly used term in the title or the abstract | 1 |
|  |  | (*b*) Provide in the abstract an informative and balanced summary of what was done and what was found | 2 |
| Introduction | | | |
| Background/rationale | 2 | Explain the scientific background and rationale for the investigation being reported | 3 |
| Objectives | 3 | State specific objectives, including any prespecified hypotheses | 3/4 |
| Methods | | | |
| Study design | 4 | Present key elements of study design early in the paper | 5 |
| Setting | 5 | Describe the setting, locations, and relevant dates, including periods of recruitment, exposure, follow-up, and data collection | 5 |
| Participants | 6 | *Cohort study*—Give the eligibility criteria, and the sources and methods of selection of participants. Describe methods of follow-up  *Case-control study*—Give the eligibility criteria, and the sources and methods of case ascertainment and control selection. Give the rationale for the choice of cases and controls  *Cross-sectional study*—Give the eligibility criteria, and the sources and methods of selection of participants | 5 |
| Variables | 7 | Clearly define all outcomes, exposures, predictors, potential confounders, and effect modifiers. Give diagnostic criteria, if applicable | 5/6 |
| Data sources/ measurement | 8* | For each variable of interest, give sources of data and details of methods of assessment (measurement). Describe comparability of assessment methods if there is more than one group | 6 and suppl. |
| Bias | 9 | Describe any efforts to address potential sources of bias | 7 |
| Study size | 10 | Explain how the study size was arrived at | 5 |
| Quantitative variables | 11 | Explain how quantitative variables were handled in the analyses. If applicable, describe which groupings were chosen and why | 5/6 |
| Statistical methods | 12 | (*a*) Describe all statistical methods, including those used to control for confounding | 7/8 |
|  |  | (*b*) Describe any methods used to examine subgroups and interactions |  |
|  |  | (*c*) Explain how missing data were addressed |  |
|  |  | (*d*) *Cohort study*—If applicable, explain how loss to follow-up was addressed  *Case-control study*—If applicable, explain how matching of cases and controls was addressed  *Cross-sectional study*—If applicable, describe analytical methods taking account of sampling strategy |  |
|  |  | (*e*) Describe any sensitivity analyses |  |
| **Results** | | | |
| Participants | 13* | (a) Report numbers of individuals at each stage of study—eg numbers potentially eligible, examined for eligibility, confirmed eligible, included in the study, completing follow-up, and analysed | 8 |
|  |  | (b) Give reasons for non-participation at each stage |  |
|  |  | (c) Consider use of a flow diagram |  |
| Descriptive data | 14* | (a) Give characteristics of study participants (eg demographic, clinical, social) and information on exposures and potential confounders | 8 (+table11) |
|  |  | (b) Indicate number of participants with missing data for each variable of interest |  |
|  |  | (c) *Cohort study*—Summarise follow-up time (eg, average and total amount) |  |
| Outcome data | 15* | *Cohort study*—Report numbers of outcome events or summary measures over time | 8 – 11  (+table2) |
|  |  | *Case-control study—*Report numbers in each exposure category, or summary measures of exposure |  |
|  |  | *Cross-sectional study—*Report numbers of outcome events or summary measures |  |
| Main results | 16 | (*a*) Give unadjusted estimates and, if applicable, confounder-adjusted estimates and their precision (eg, 95% confidence interval). Make clear which confounders were adjusted for and why they were included | 8 – 11  (+table2) |
|  |  | (*b*) Report category boundaries when continuous variables were categorized |  |
|  |  | (*c*) If relevant, consider translating estimates of relative risk into absolute risk for a meaningful time period |  |
| Other analyses | 17 | Report other analyses done—eg analyses of subgroups and interactions, and sensitivity analyses | 11 |
| **Discussion** | | | |
| Key results | 18 | Summarise key results with reference to study objectives | 12 |
| Limitations | 19 | Discuss limitations of the study, taking into account sources of potential bias or imprecision. Discuss both direction and magnitude of any potential bias | 16 |
| Interpretation | 20 | Give a cautious overall interpretation of results considering objectives, limitations, multiplicity of analyses, results from similar studies, and other relevant evidence | 12 - 16 |
| Generalisability | 21 | Discuss the generalisability (external validity) of the study results | 16 |
| **Other information** | | | |
| Funding | 22 | Give the source of funding and the role of the funders for the present study and, if applicable, for the original study on which the present article is based | 16/17 |

### **SUPPLEMENTARY DISCUSSION**

**Figure 3. Acylcarnitine production and main roles in mitochondrial function.**

**Legend:** Abbreviations: CIT, cytosol; MIT, mitochondria; PEX, peroxisome; LC, long-chain; MC, medium-chain; SC, short-chain; CoA, coenzyme A; LACS, long-chain acyl-coenzyme A synthetase; CACT, carnitine/acylcarnitine translocase; CPT1 and CPT2, carnitine palmitoyl-transferase 1 and 2; CRAT, carnitine O-acetyltransferase; ATP, adenosine triphosphate; BCAA, branched-chain amino acid; OC Fatty Acid, Odd-chain fatty acid. Model based on Dambrova et al. (2022)^[3]^ and Li et al. (2019)^[4]^.

The main function of ACs is involved in long-chain fatty acids β-oxidation, where they are carriers transporting long-chain Fatty Acids into mitochondria after their activation by the link with coenzyme A via LACS. Long- and medium-chain acyl-CoAs are converted into LC Acylcarnines by CPT1 which is located on the outer mitochondrial membrane. Under catalysis of CACT, these LC Acylcarnines are imported through the mitochondrial membranes into the mitochondrial matrix (to be noted, part of the ACs involved are produced in peroxisomes). Then they are converted back to the corresponding long-/medium-chain acyl-CoAs by CPT2. These products will participate to β-oxidation for the production of ATP. The end products, acetyl-CoAs, are converted to acetylcarnitines (C2) by carnitine O-acetyltransferase (CRAT), which regulates the acetyl-CoA/free CoA ratio to prevent depletion of free CoA. Also, branched-chain amino acid (BCAA) and odd-chain fatty acids metabolization produce propionyl-carnitine (C3) which allows the creation of propionyl-CoA, substrate for the Krebs cycle that generate energy through the complete oxidation of acetyl groups derived from carbohydrates, fats, and proteins.
